# Genomic epidemiologic assessment implicates prolonged silent carriage, virulence factors and transmission between staff and patients in a NICU outbreak of MRSA

**DOI:** 10.1101/2021.07.28.21261307

**Authors:** Sharline Madera, Nicole McNeil, Paula Hayakawa Serpa, Jack Kamm, Christy Pak, Carolyn Caughell, Amy Nichols, David Dynerman, Lucy M. Li, Estella Sanchez-Guerrero, Maira Phelps, Angela M. Detweiler, Norma Neff, Helen Reyes, Steve Miller, Deborah Yokoe, Joseph L. DeRisi, Lynn Ramirez-Avila, Charles R. Langelier

## Abstract

**Introduction:** Methicillin-resistant *Staphylococcus aureus* (MRSA) is an important pathogen in neonatal intensive care units (NICU) that carries significant morbidity and mortality. Improving our understanding of MRSA transmission dynamics, especially among high risk patients, is an infection prevention priority.

**Methods:** We investigated a cluster of clinical MRSA cases in the NICU using a combination of epidemiologic review and whole genome sequencing (WGS) of isolates from clinical and surveillance cultures obtained from patients and healthcare personnel (HCP).

**Results:** Phylogenetic analysis identified two genetically distinct phylogenetic clades and revealed multiple silent transmission events between HCP and infants. The predominant outbreak strain harbored multiple virulence factors. Epidemiologic investigation and genomic analysis identified a HCP colonized with the dominant MRSA outbreak strain who cared for the majority of NICU patients who were infected or colonized with the same strain, including one NICU patient with severe infection seven months before the described outbreak. These results guided implementation of infection prevention interventions that prevented further transmission events.

**Conclusion:** Silent transmission of MRSA between HCP and NICU patients likely contributed to a NICU outbreak involving a virulent MRSA strain. WGS enabled data-driven decision making to inform implementation of infection control policies that mitigated the outbreak. Prospective WGS coupled with epidemiologic analysis can be used to detect transmission events and prompt early implementation of control strategies.

**Summary:** Whole genome sequencing identified a NICU outbreak of MRSA preceded by silent transmission between health care personnel and patients seven months prior to outbreak detection. This study suggests prospective genomic analysis may identify isolates with high virulence potential, offering an early opportunity for intervention.

## INTRODUCTION

*Staphylococcus aureus* (*S. aureus)* remains a leading cause of hospital-acquired infections in the neonatal intensive care unit (NICU)[1]. Critically-ill and preterm neonates are particularly vulnerable to invasive infections with methicillin-resistant *S. aureus* (MRSA), which confers significant morbidity, mortality [2,3] and financial costs[3]. Despite this, the risk factors and mechanisms of transmission of MRSA in NICUs are incompletely understood.

While MRSA colonization has been identified as a risk factor for the development of invasive infection[4], the specific factors contributing to transmission of MRSA between NICU patients are not well defined. Factors previously identified include environmental reservoirs, as outbreak-associated MRSA strains can exhibit longer environmental persistence than sporadic-MRSA isolates[5]. Parental colonization has been described as a source of MRSA both via vertical[6] and horizontal[7,8] transmission; however, these transmission events are usually self-limited and have not been implicated in outbreaks. Exposure to colonized health-care personnel (HCP) or patients may play a role in precipitating outbreaks. In many hospitals, patients are tested for MRSA nares colonization upon admission, however HCP generally are not [9–11].

Here we report an outbreak investigation of MRSA in a NICU in which HCP were identified as a potential key link in transmission and colonization. Whole genome sequencing (WGS) further identified circulation of a highly virulent outbreak strain for at least 7 months prior to outbreak recognition in the NICU. We highlight an opportunity for prospective WGS in the surveillance setting to aid in the early identification of predominant circulating strains whose virulence may facilitate spread and invasive infection.

## METHODS

### Setting

The University of California San Francisco (UCSF) NICU contains 58 beds and is located within the 183-bed UCSF Benioff Children’s Hospital.

### Ethics Statement

This study was approved by the UCSF Institutional Review Board (IRB) under protocol 17-24056, which permitted WGS analysis of clinical isolates and review of clinical microbiology results and other electronic health record data.

### Microbial culture

Bacterial cultures were inoculated into BACTEC blood culture bottles (blood). MRSA surveillance cultures were performed using selective chromogenic agar (Thermo Scientific Spectra MRSA).

### Metagenomic sequencing

DNA was extracted from cultured isolates using the Zymo ZR Fungal/Bacterial DNA MiniPrep Kit, 100 ng of DNA from each sample was then sheared with NEB fragmentase and used to construct sequencing libraries using the NEBNext Ultra II Library Prep Kit (New England Biolabs). Adaptor ligated samples underwent 12 cycles of amplification with dual-indexing primers using the NEBNext Ultra II Q5 polymerase. Libraries were then quantified with Qubit and quality checked with a Bioanalyzer High Sensitivity DNA chip. Samples were pooled and sequenced on an Illumina NextSeq instrument using a NextSeq 500/500 Mid Output kit v2.5 (300 cycles).

### Bioinformatic analyses

Raw sequences were analyzed using the SNP Pipeline for Infectious Disease (SPID)[12]. SPID aligned samples against S. aureus reference genome USA300 TCH1516 using minimap2, followed by samtools to perform an mpileup. Julia code was then run to call a consensus allele at each position, and the SNP instances were computed between every pair of samples. Phylogenetic analysis was performed using randomized axelerated maximum likelihood (RAxML)[13]. Phylogenetic trees were further visualized with ETE Python API[14].

ARGannot was used to identify antimicrobial resistance genes from quality-filtered raw sequence data for each of the evaluated isolates[15]. Genes with greater than 99% gene coverage were included in the analyses. SCC mec elements were detected using SCC mec finder[16]. Multilocus sequence type (MLST) typing was performed using MLST 2.0[17]. Detection of *S. aureus* virulence genes was performed using VirulenceFinder[18].

## RESULTS

### Outbreak Description

After four infants in the NICU with invasive MRSA infections were identified in an eight-day period, an investigation was initiated. Retrospective review of microbiology data identified two additional positive MRSA cultures from infants in the same geographic zone of the NICU two months prior (Supplemental Table 1). NICU-wide surveillance screening of all infants and staff identified 16 other cases of MRSA colonization in asymptomatic infants or HCP (Supplemental Figure 1). Review of prior MRSA isolates identified an isolate from an infant with extensive MRSA infection 7 months prior (ID 10B, Supplemental Table 1). In total, 23 MRSA isolates from 18 patients and 5 HCP were evaluated, and baseline characteristics of each subject are listed in Supplemental Table 1. Antimicrobial susceptibility testing (AST) demonstrated similarities across some isolates, suggestive of an outbreak (Supplemental Figure 1).

### Genomic Analyses

All isolates underwent WGS. Assembly by alignment against the chromosomal sequence of the National Center for Biotechnology Information (NCBI) reference strain *S. aureus* USA300 TCH1516 confirmed an outbreak composed of two genetically distinct clades (Figure 1A). Pairwise single nucleotide polymorphism (SNP) was used as a measure of genetic relatedness between isolates. A cutoff of 23 SNPs was used to define isolate belonging to the same *S. aureus* phylogenetic clade[19]. Phylogenetic analysis revealed and outbreak composed of two distinct phylogenetic clades; clade 1 was composed of 10 infants and one HCP (11), and clade 2 composed of three infants and 1 HCP (15, Figure 1A). 8 of the 23 cases were deemed unrelated to either of the two clades based on SNP distance.

**Figure 1.**
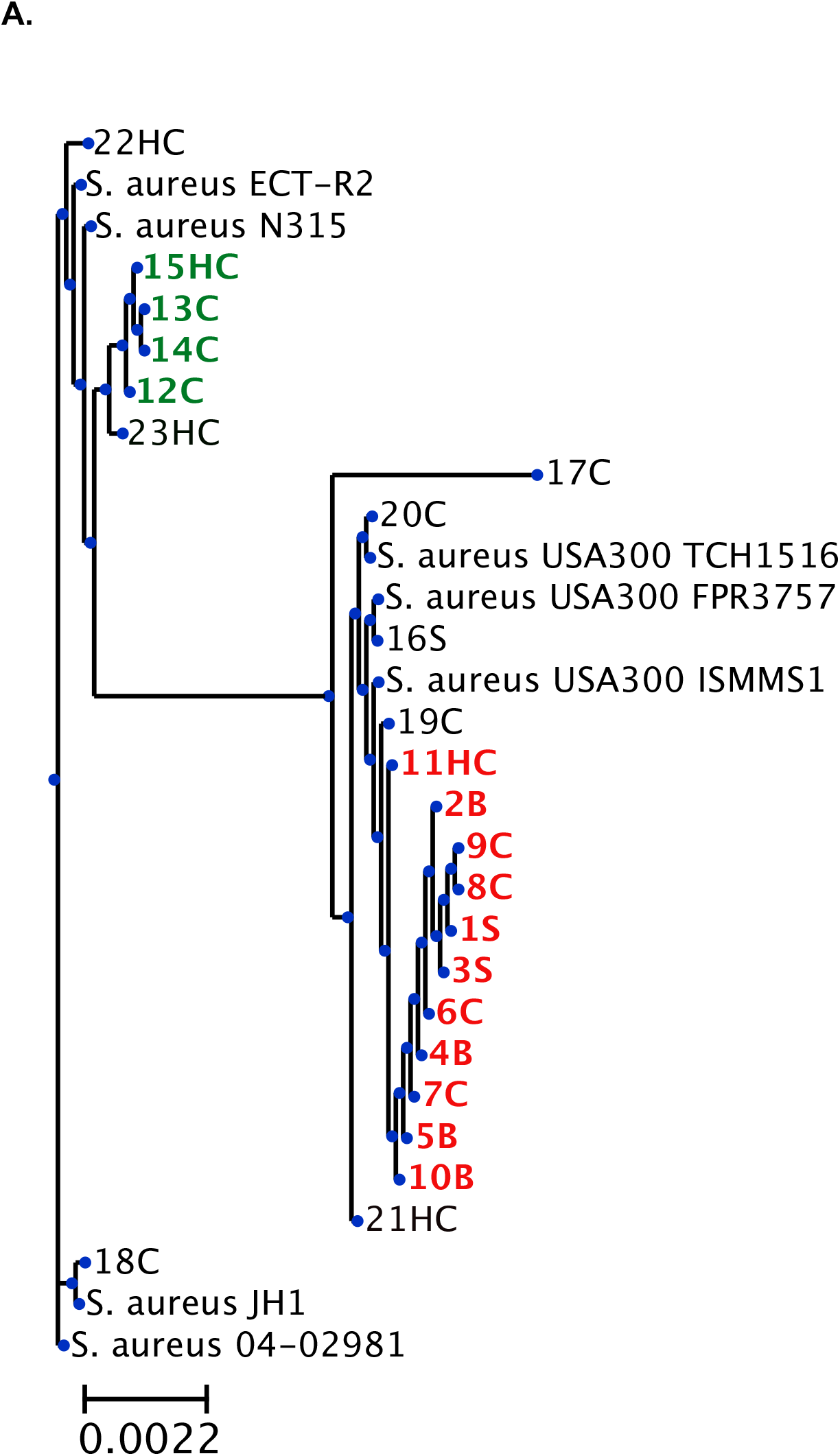
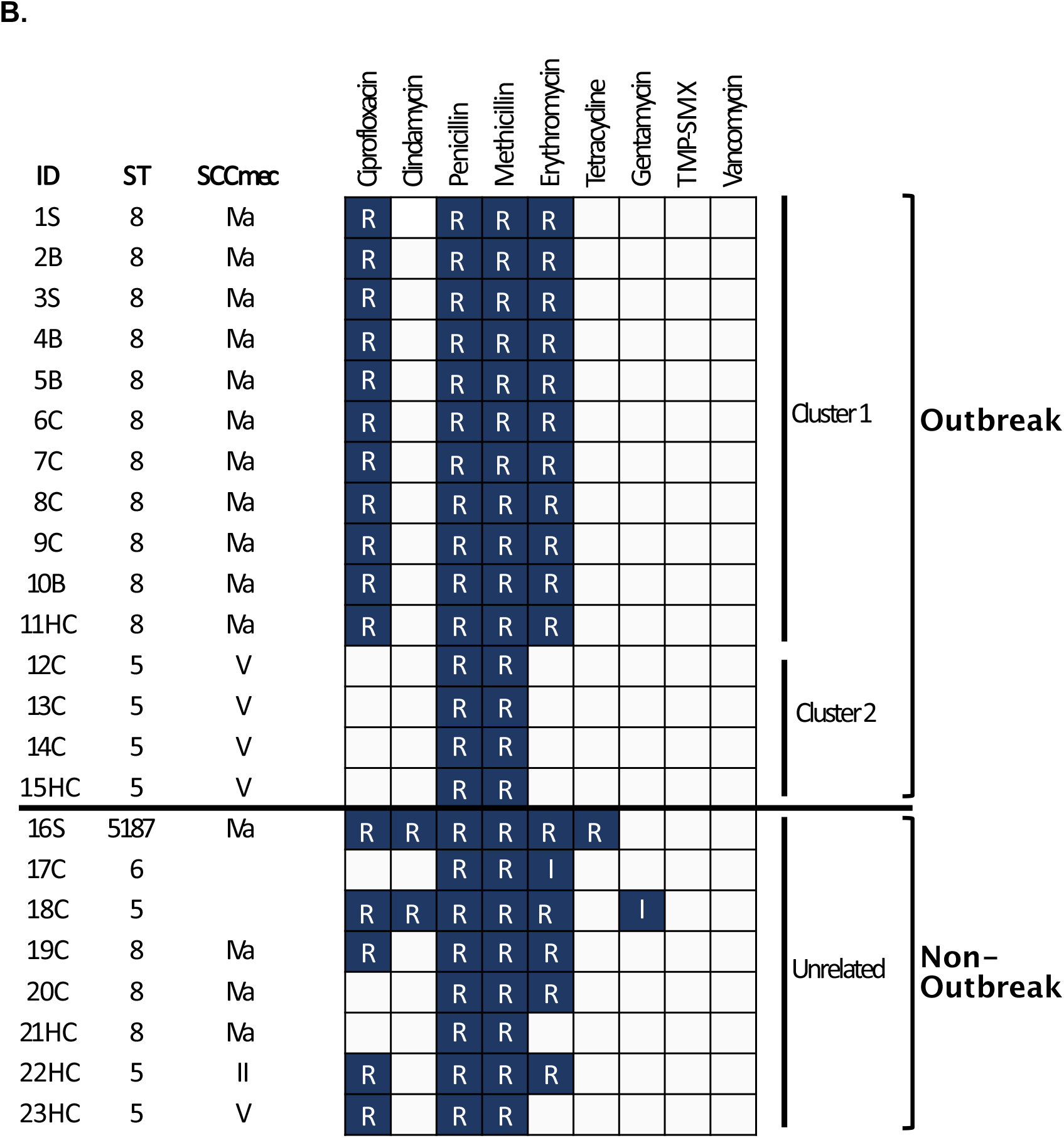
Phylogenetic Analysis and Phenotypic Antibiogram of all Sequenced MRSA Cases. Isolates are identified by case number followed by H for healthcare personnel, B for bacteremia, S for skin and soft tissue infection, and C for colonization. (A) Cases associated with clade 1 outbreak strain labeled red, and clade 2 outbreak strain labeled green. Reference *S. aureus* strains ECT-R2, N315, JH1, 04-02981, and USA 300 strains TCH1516, FPR3757, and ISMMS1, are shown. (B) Antimicrobial susceptibility pattern for all cases sequenced. Susceptibility to antibiotics are denoted as resistant (R) or intermediate (I). Sequence type (MLST) and Staphylococcal Cassette Chromosome (SCC) mec type are shown.

Clade 1 exhibited marked genomic similarity between the 11 isolates that make up the clade with a median SNP difference of 3, range 0-6. Similarly, four isolates comprising clade 2 shared striking genetic similarity with a mean SNP difference of 7, range 0-23. An isolate belonging to case ID 23EC was approximately 800 SNPs away from clade 2, and thus not likely part of a singular transmission event. Interestingly, clade 1 samples were more closely related to ST8 MRSA reference strains USA300 TCH 1516, ISMMS1, and FPR3757, with approximately 140 SNP differences; while clade 2 was more closely related to ST5 MRSA reference strain N315 with an approximately 700 SNP difference. MRSA reference strain N315 harbored a 17,000 SNP difference from clade 1, and ST8 MRSA reference strains carried roughly a 20,000 SNP difference from clade 2.

Confirmation of a highly related cluster of MRSA isolates through WGS prompted further epidemiologic investigation that revealed that eight out of the eleven infants associated outbreak clade 1 had been cared for by HCP 11 (Figure 2). HCP 11 also cared for patient 10B, who was identified 7 months prior to clade 1, raising the possibility that HCP 11 could be the source of the current outbreak. Almost all patients associated with the outbreak strains were born prematurely and exhibited low birthweight (Supplemental Table 1). None of the cases associated with outbreak clade 2 group exhibited active infection.

**Figure 2.**
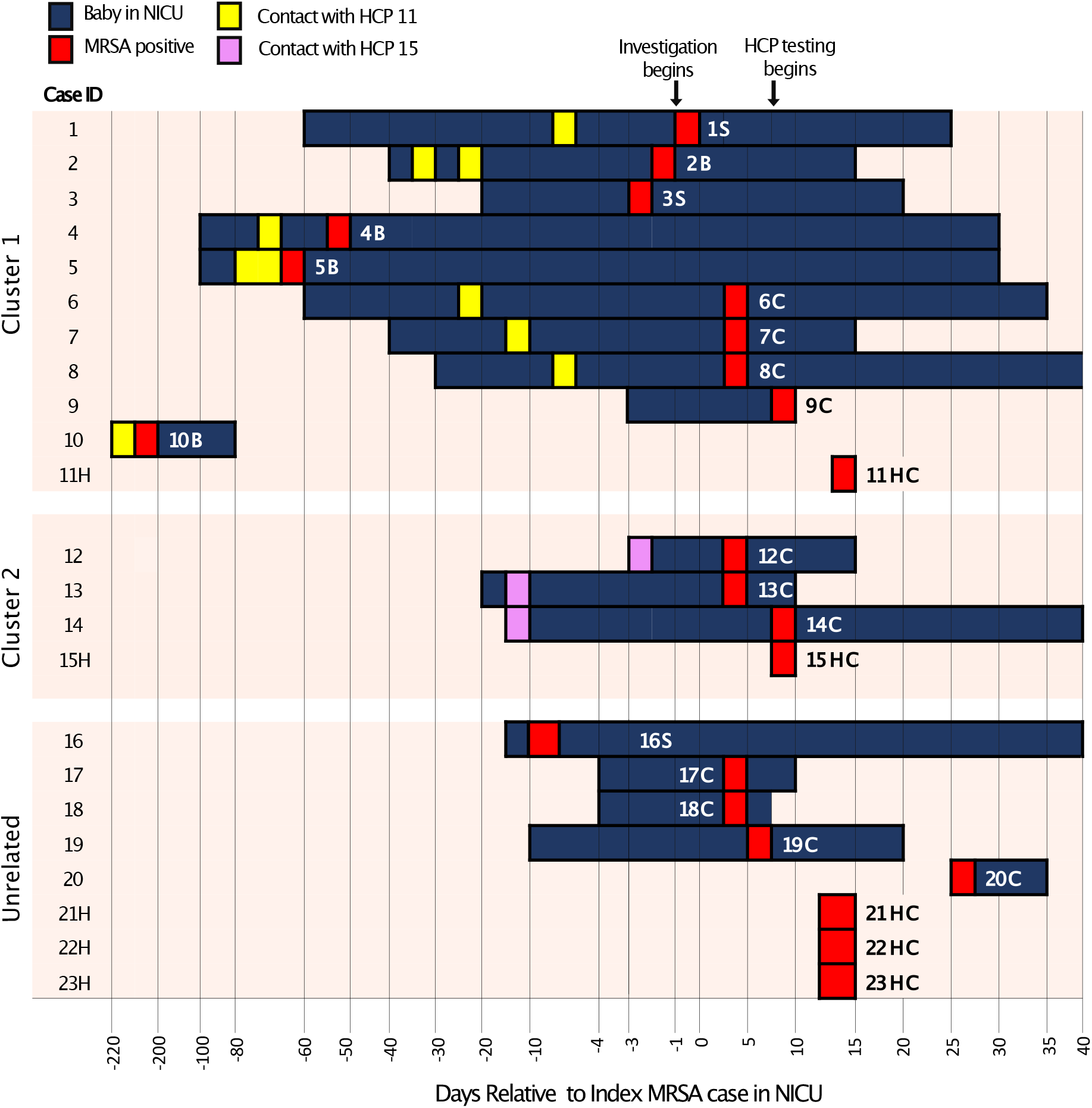
Epidemiologic Tracing of MRSA Outbreak in the NICU. Index case and 22 other cases associated with an MRSA outbreak. Healthcare personnel (HCP) are indicated by an H. Epidemiologic link to each of two HCP iis ndicated by yellow (11H) and purple (15H), respectively.

*In silico* MLST typing revealed that outbreak clade 1 was sequence type (ST)-8, and smaller outbreak clade 2 was ST-5 (Figure 1B). Staphylococcal cassette chromosome (SCC) *mec* typing revealed further similarities among outbreak clades, with clade 1 samples identified to harbor SCC*mec* IVa, and clade 2 cases identified to harbor SCC*mec* V (Figure 1B). Notably, MRSA isolate belonging to case ID 19C shared identical AST, MLST and SCCmec typing as clade 1 isolates but was approximately 270 SNPs away from clade 1, and therefore distinct from the predominant outbreak clade.

We next carried out antimicrobial-resistance, virulence and toxin gene assessment. Cases associated with both outbreak clade 1 and clade 2 exhibited similar antimicrobial-resistance gene profiles (Figure 3) and concordant phenotypic antimicrobial-resistance profile (Figure 1B). Interestingly, across non-outbreak associated isolates, isolates 17C and 18C, were found to lack the *mecA* gene, associated with resistance to beta-lactam antibiotics like methicillin and penicillin, despite being phenotypically resistant. Further, 18C also lacked the *blaZ* gene, associated with resistance to penicillin, despite being phenotypically resistant. Harboring of aminoglycoside resistance genes, *sat4A* or *spc*, was not associated with phenotypic antimicrobial-resistance.

**Figure 3.**
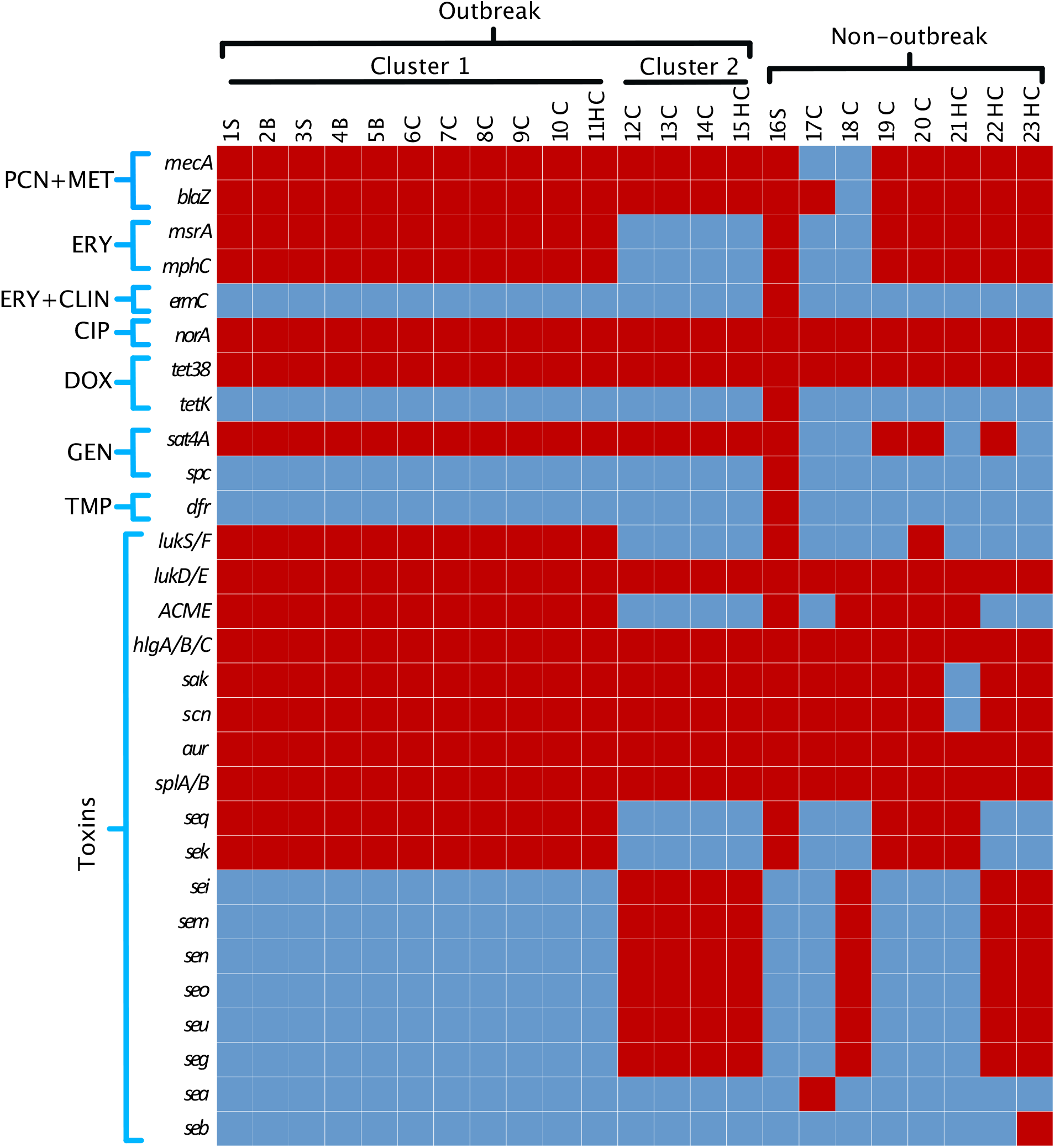
Antimicrobial resistance and toxin genes identified by WGS of MRSA isolates. Genes associated with antibiotic resistance and toxins are shown for all cases sequenced. Gene presence is depicted in red and gene absence is depicted in blue. PCN denotes penicillin, MET methicillin, ERY erythromycin, CLIN clindamycin, CIP ciprofloxacin, DOX doxycycline, GEN gentamycin, and TMP trimethoprim. Genes associated with toxins are shown.

Presence of genes encoding virulence factors or toxins revealed similarities across clade 1 and clade 2 isolates. Interestingly, clade 1 isolates harbored *lukS/F*, which encodes the cytotoxin Panton-Valentine leukocidin (PVL), and the arginine catabolic mobile element (ACME), which are associated with severe invasive infection**[20]** or the promotion of persistence and skin-colonization**[21]**, respectively. Clade 1 was also found to harbor superantigen enterotoxin genes *seq* and *sek***[22]**. In further contrast to clade 1, clade 2 isolates contained the enterotoxin gene clade *sei, sem, sen, seo, seg*, and *seu* encoded by superantigen enterotoxin gene clade *egc***[23,24]**. All isolates were found to contain the gamma-hemolysin genes *hlgA/B/C* and the leukocidin genes *lukD/E*.

## DISCUSSION

Real-time deployment of WGS provided high resolution assessment of an outbreak involving dynamic exchange of genetically distinct MRSA strains between HCPs and patients in a NICU. Phylogenetic analysis uncovered two outbreak clades in the NICU, both involving HCP who had contact with most, if not all, clade associated isolates. Clade 1 was associated with invasive and severe infections, while clade 2 was associated with asymptomatic carriage.

Surprisingly, outbreak clade 1 included patient 10B, who was admitted to the NICU 7 months prior to the outbreak and was cared for by HCP 11, also found to be colonized by clade 1. This raises the possibility that HCP 11 may have played a role in the dissemination of the outbreak clade 1. Not all clade 1 isolates had an epidemiologic link to HCP 11, however, which raises the possibility of unrecognized inter-HCP transmission of the clade 1 strain or, alternatively, the presence of an environmental reservoir.

Assessment of *S. aureus* virulence factors in the strains from each clade suggested potential molecular determinants driving outbreak emergence. Clade 1 isolates associated with the pandemic clonal lineage USA300, classically regarded as a community-associated (CA-) MRSA strain. Clade 2 isolates most closely associated with strain NS315, a hospital-associated (HA-) MRSA strain. Both outbreak isolates harbored the lukS/F gene that encodes for the cytotoxin Panton-Valentine leukocidin (PVL), a protein first associated with epidemic CA-MRSA strains that are more likely to cause sepsis, necrotizing pneumonia, and necrotizing fasciitis[20]. However, unlike clade 2, the clade 1 strain harbored the genomic island referred to as ACME. First described in CA-MRSA strain USA300 in 2006, ACME likely originated from *S. epidermidis*[25]. This element is composed of 33 putative genes and two operons, *arc* and *opp*.

The arc operon encodes genes thought to be important in arginine catabolism and have been shown to confer increased survival in the acidic skin environment[26]. The AMCE *speG* gene, which encodes a spermidine acetyltransferase, has been shown to facilitate survival in the presence of lethal polyamines, spermidine and spermine that are produced by human skin[26]. Therefore, ACME-containing *S. aureus* are poised to execute epidemic, severe infections, likely explaining why clade 1 isolate was able to circulate in the NICU for over nine months, causing multiple episodes of severe infection along the way. Rapid identification of potentially high-virulence strains highlights an opportunity for preventative surveillance of this high-risk population and the HCP who provide their care.

Notably, isolates 16P and 20C also harbored the *lukS/F* gene and the ACME element. These isolates were not associated with a clade, and likely reflect singular acquisitions of highly virulent strains. Clade 1 and clade 2 isolates, on the other hand, had HCP to provide an epidemiologic link that could suggest repeated exposures to the highly virulent isolates that led to the pathologic infection and/or colonization of numerous neonates. This highlights the possibility that silent transmission of antimicrobial resistant organisms is taking place throughout the healthcare system, leaving our most vulnerable patients at greatest risk.

Genomic analysis also provided insight regarding antimicrobial resistance mechanisms of the evaluated isolates and revealed an unexpected discordance between antimicrobial resistance genes and phenotypic antimicrobial resistance. Resistance to methicillin and other β-lactam antibiotics in *S. aureus* is largely mediated by production of an altered penicillin binding protein (PBP)2a, which is encoded by *mecA* and is harbored in the large genetic element designated SCC*mec*[27]. Presence of *mecA* is considered hallmark in the identification of MRSA. Interestingly, infant isolates 17C and 18C did not harbor *mecA* (Figure 3), despite still exhibiting resistance to methicillin (Figure 1B). Borderline and low-level methicillin resistance (MIC 2-4 mg/L) in *mecA* deficient MRSA has been ascribed to overproduction of a penicillinase[28] or alteration in PBP expression[29]. Infant isolate 18C was further noted to lack the *blaZ* gene, but still exhibited phenotypic penicillin resistance. Nonetheless, this highlights the presence of alternative mechanisms of antibiotic resistance, which merit further investigation into alternative genetic determinants related to resistance.

Control of both outbreak clades was obtained through several measures including temporary closure of the NICU to new admissions, global contact isolation, positive case cohorting, reinforcement of hand hygiene and PPE practices, extensive environmental disinfection, and patient, parent, and HCP decolonization. All MRSA colonized patients and HCP underwent MRSA decolonization with nasal mupirocin (patients) or nasal povidone iodine (HCP) and topical chlorhexidine if they were eligible. Parents of MRSA colonized patients were offered decolonization with nasal povidone iodine and topical chlorhexidine. Ongoing prospective surveillance in the NICU has not identified further clade 1 or 2 associated MRSA isolates 6 months after the outbreak investigation, underscoring the importance and efficacy of decolonization, as has been noted in the case of parental reservoirs[30].

As evidenced in this work and others[31], conventional methods of identifying pathogens and outbreak investigation, which have largely relied on MLST[32], and prior to that, Pulse-field Gel Electrophoresis (PFGE)[10], lack the resolution necessary to identify the source and transmission of an outbreak. For instance, isolate belonging to ID 19C shared identical MLST typing as clade 1, as well as the same AST and SSC*mec* typing. The added resolution of WGS allowed us to accurately identify outbreak clades and infer direction of transmission, which proved pivotal in mitigating the outbreak.

Although the directionality of MRSA transmission cannot be confirmed, both outbreak clades identified were associated with HCP through genetic and epidemiologic linkage. Prior MRSA outbreak studies in the NICU have also identified the potential contribution of the HCP reservoir[10,33], but few, if any, have shown prolonged circulation in the hospital environment through silent transmission events that have led to significant patient morbidity, as this work demonstrates. The high risk of ongoing transmission events originating from undetected MRSA carriers suggests that increasing the frequency of surveillance efforts through genomic diagnostics may result in decreased transmission. This begs the consideration of prospective genomic surveillance that includes the HCP reservoir as a way to identify uniquely virulent MRSA strains with theoretical high epidemic potential (PVL and ACME carriage, etc). While economic modeling studies of prospective genomic MRSA surveillance suggests it’s cost-effectiveness[34], it may still pose various difficulties such as when to include HCP during prospective screening and who should be included, at what intervals should screening occur, and should clinical triggers be used to initiate HCP screening, among others[35]. More research into economic and clinical benefits of HCP inclusion in prospective genomic surveillance is necessary.

This study has limitations that should be considered. While this work highlights potential that silent transmission of MRSA may occur for months before detection of an outbreak, we were unable to sequence all MRSA isolates detected in the NICU during the seven-month gap between sample the index case and the outbreak, nor isolates prior to the index case. This limits our ability to definitively pinpoint the introduction and complex transmission dynamics of outbreak clade 1. Further, the source of introduction of clade 1 and clade 2 remain elusive and necessitates consideration of other potential reservoirs besides the affected HCPs, such as other HCPs, parents, and the environment. However, environmental samples were unavailable. Nevertheless, the high degree of genetic similarity of clade 1 isolates, evidenced by low SNP difference, suggest clade 1 isolates were part of a singular transmission event and thus highlight the dangers of silent transmission of highly virulent isolates can pose. Finally, we were not able to validate the discrepancy between AST and *mecA* exhibited by isolates belonging to ID 17C and 18C.

In summary, our results suggest that transmission between HCP and patients contributed to a NICU outbreak involving a uniquely virulent MRSA strain. WGS enabled data-driven implementation of infection prevention strategies that prevented transmission of the outbreak strains and identified a potential source. Prospective genomic epidemiology of hospital acquired infections may help identify occult transmission events that may precede outbreaks of MRSA and other pathogens. Our findings corroborate prior work demonstrating the utility of WGS in outbreak management[18,36,37]. Given the increasing availability and affordability of WGS, genomic epidemiology of CDC priority pathogens from staff and patients may be a useful measure for identifying early colonization and silent transmission events of virulent strains prior to outbreak emergence.

## Data Availability

Data is available upon request.NHLBI K23HL138461-01A1 (CL)
NIH 5T32AI007641-19 (SM)

