## Supplemental Figure 1 for "Genomic epidemiologic assessment implicates prolonged silent carriage, virulence factors and transmission between staff and patients in a NICU outbreak of MRSA"

| ID | Ciprofloxacin | Clindamycin | Penicillin | Methicillin | Erythromycin | Tetracycline | Gentamycin | TMP-SMX | Vancomycin |
|---|---|---|---|---|---|---|---|---|---|
| 1P | R | | R | R | R | | | | |
| 2B | R | | R | R | R | | | | |
| 3P | R | | R | R | R | | | | |
| 4B | R | | R | R | R | | | | |
| 5B | R | | R | R | R | | | | |
| 6C | R | | R | R | R | | | | |
| 7C | R | | R | R | R | | | | |
| 8C | R | | R | R | R | | | | |
| 9C | R | | R | R | R | | | | |
| 10B | R | | R | R | R | | | | |
| 11HC | R | | R | R | R | | | | |
| 12C | | | R | R | | | | | |
| 13C | | | R | R | | | | | |
| 14C | | | R | R | | | | | |
| 15HC | | | R | R | | | | | |
| 16P | R | R | R | R | R | R | | | |
| 17C | | | R | R | I | | | | |
| 18C | R | R | R | R | R | | I | | |
| 19C | R | | R | R | R | | | | |
| 20C | | | R | R | R | | | | |
| 21HC | | | R | R | | | | | |
| 22HC | R | | R | R | R | | | | |
| 23HC | R | | R | R | | | | | |

**Supplemental Figure 1. Antimicrobial Susceptibility Testing.** Antimicrobial susceptibility pattern for all cases sequenced. Cases from healthcare providers (H), bacteremia (B), skin and soft tissue infection (S), or colonization (C) are indicated with the case ID. Susceptibility to antibiotics are denoted as resistant (R) or intermediate (I).
