## Supplemental Table 1 for "Genomic epidemiologic assessment implicates prolonged silent carriage, virulence factors and transmission between staff and patients in a NICU outbreak of MRSA"

| ID | First Positive Culture* | Sex | GA at birth | Birthweight (g) | Reason for ICU Admission | MRSA Manifestation | Site |
|---|---|---|---|---|---|---|---|
| 1 | 0 | Female | 25w0d | 800 | Prematurity, RDS | SSTI† | Wound |
| 2 | -1 | Female | 32w5d | 1840 | Prematurity, Scimitar Syndrome, Aortic Coarctation, VSD, pHTN | Bacteremia | Blood |
| 3 | -3 | Male | 33w1d | 2175 | Prematurity, RDS, Congenital Syphilis | SSTI† | Wound |
| 4 | -55 | Male | 27w1d | 1395 | Prematurity, RDS, PDA | Bacteremia | Blood |
| 5 | -65 | Male | 27w1d | 935 | Prematurity, RDS, PDA, PFO | Bacteremia | Blood |
| 6 | 3 | Female | 32w1d | 1420 | Prematurity, Omphalocele | Colonization | Nasal |
| 7 | 3 | Male | 33w0d | 2760 | Prematurity, Myelomeningocele, | Colonization | Nasal |
| 8 | 3 | Male | 26w0d | 830 | Prematurity, Perforated Viscus | Colonization | Nasal |
| 9 | 8 | Male | 28w0d | 1300 | Prematurity, RDS | Colonization | Nasal |
| 10 | -220 | Female | 29w6d | 1515 | Prematurity, RDS | Bacteremia, Meningitis | Blood |
| 11H | 15 | | | | | Colonization | Nasal |
| 12 | 3 | Male | 26w0d | 1675 | Prematurity, RDS | Colonization | Nasal |
| 13 | 3 | Male | 28w0d | 1330 | Prematurity, RDS | Colonization | Nasal |
| 14 | 8 | Male | 28w6d | 1030 | Prematurity, RDS | Colonization | Nasal |
| 15H | 10 | | | | | Colonization | Nasal |
| 16 | -8 | Male | 31w6d | 1572 | Prematurity, RDS | SSTI | Wound |
| 17 | 3 | Male | 40w0d | 3485 | PPHN | Colonization | Nasal |
| 18 | 3 | Male | 30w1d | 1700 | Prematurity, RDS, HIE | Colonization | Nasal |
| 19 | 5 | Female | 39w1d | 3845 | dTGA, VSD | Colonization | Nasal |
| 20 | 26 | Male | 39w2d | 3840 | Tetralogy of Fallot | Colonization | Nasal |
| 21H | 15 | | | | | Colonization | Nasal |
| 22H | 15 | | | | | Colonization | Nasal |
| 23H | 16 | | | | | Colonization | Nasal |

* days relative to index MRSA case in the NICU
† skin and soft tissue infection

**Supplemental Table 1. Characteristics of study participants.** Isolates are identified by case number followed by H for healthcare personnel (HCP). Gestational age (GA) at birth, and birthweight in grams are shown. Each patient's reason for admission to the ICU, as well as the manifestation of MRSA infection and body site of sampling is noted.
